## Supplementary Figures S1-S9 for "RNA sequencing of whole blood defines the signature of high intensity exercise at altitude in elite speed skaters"

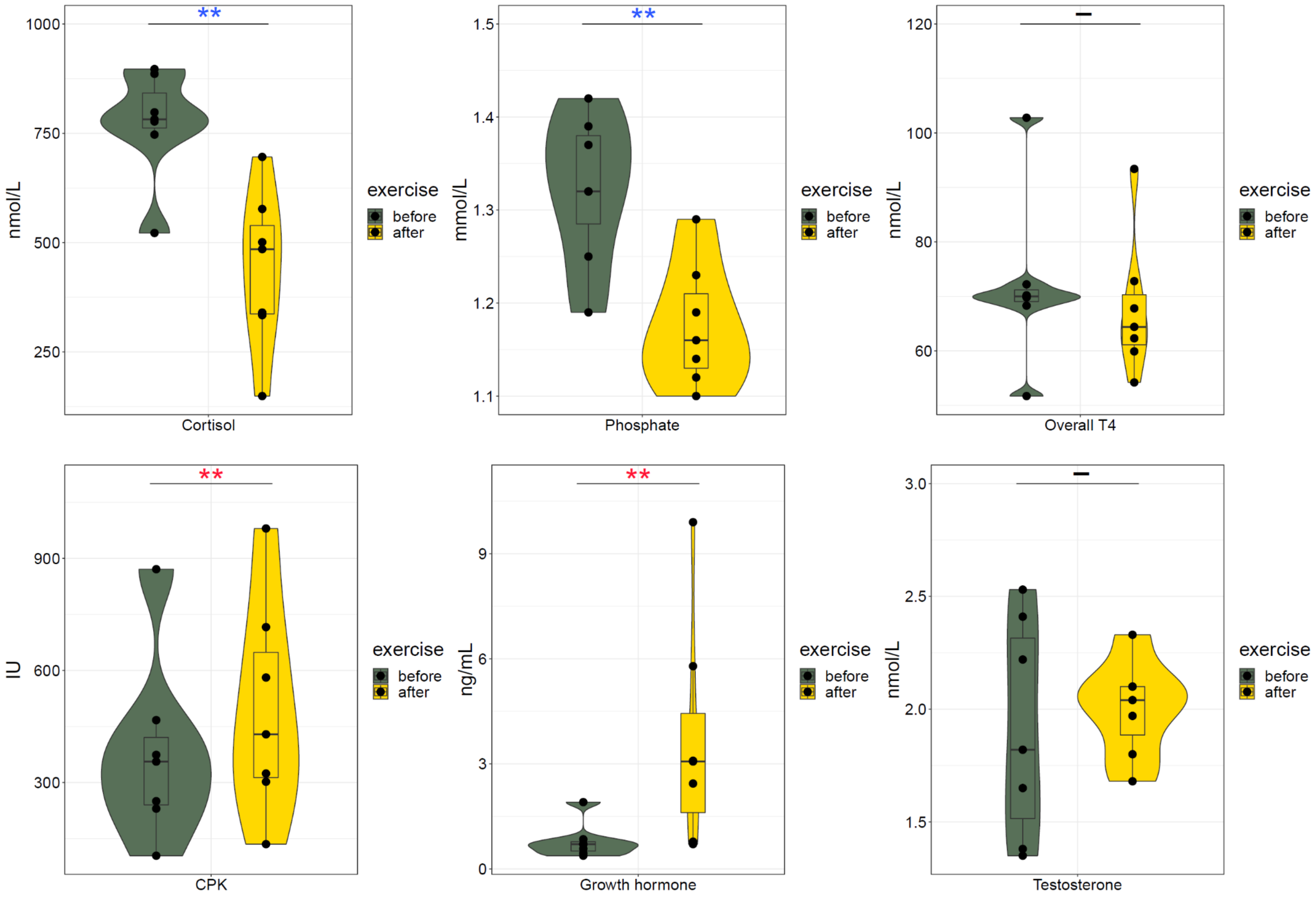


Figure S1. Blood tests taken immediately before and after the exercise bout on day 18. Full table of all performed blood tests is given in Supplementary Table S2. Paired one-sided Wilcoxon test was used to measure the significance of each change.

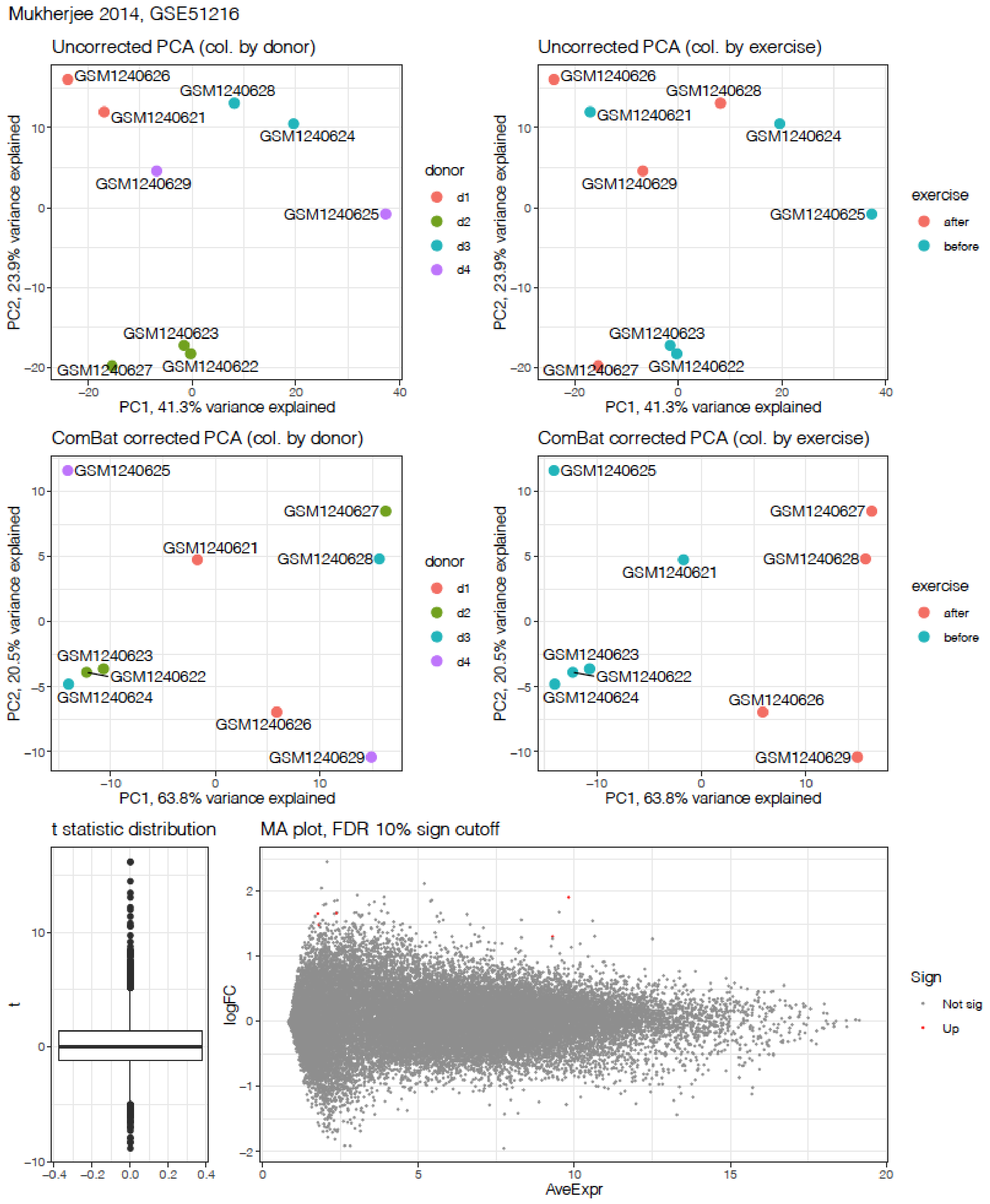


Figure S2. Microarray comparison of 4 adult athletes before and after exercise (Mukherjee 2014).

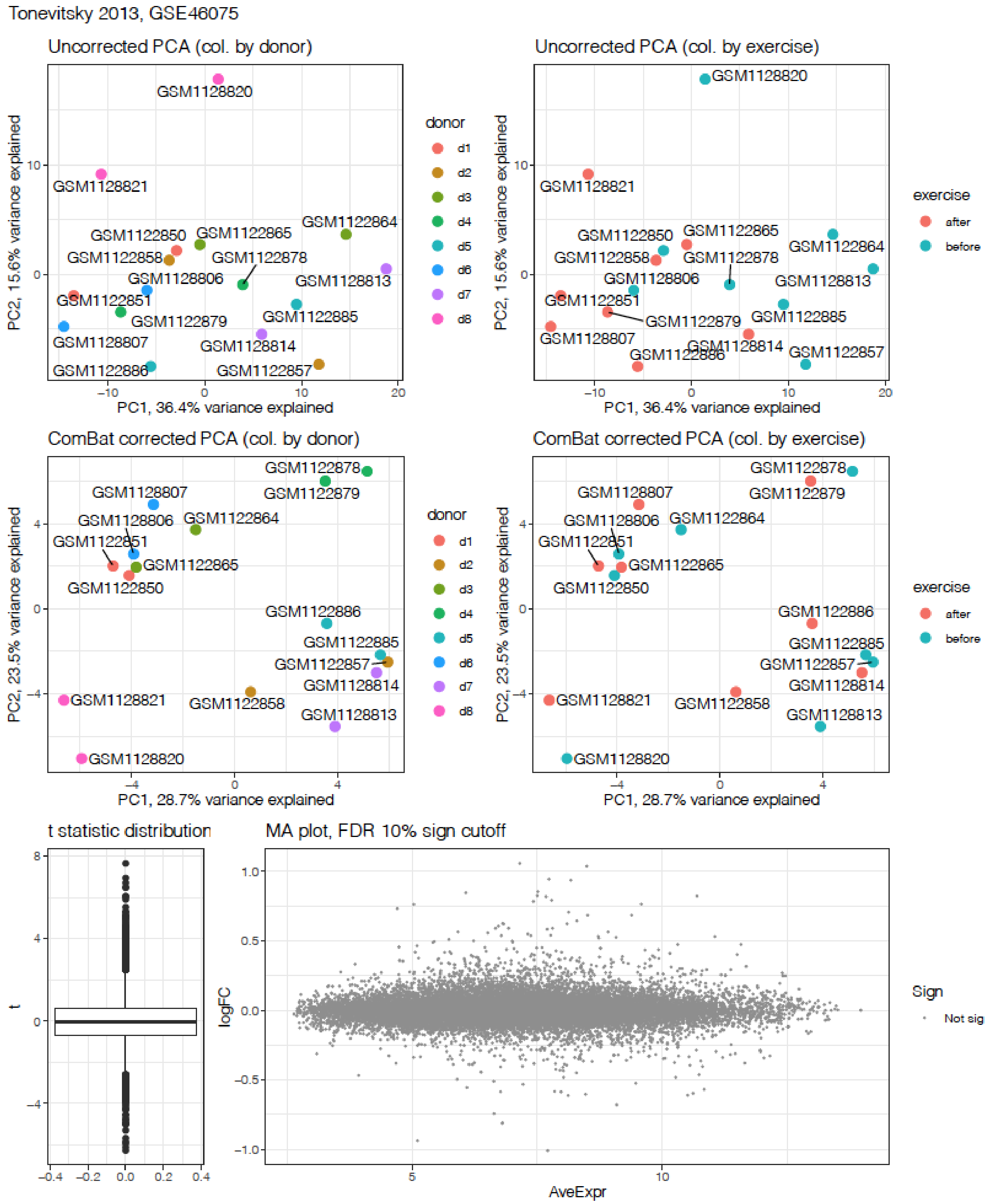


Figure S3. Microarray comparison of 8 adult athletes before and after exercise (Tonevitsky 2013).

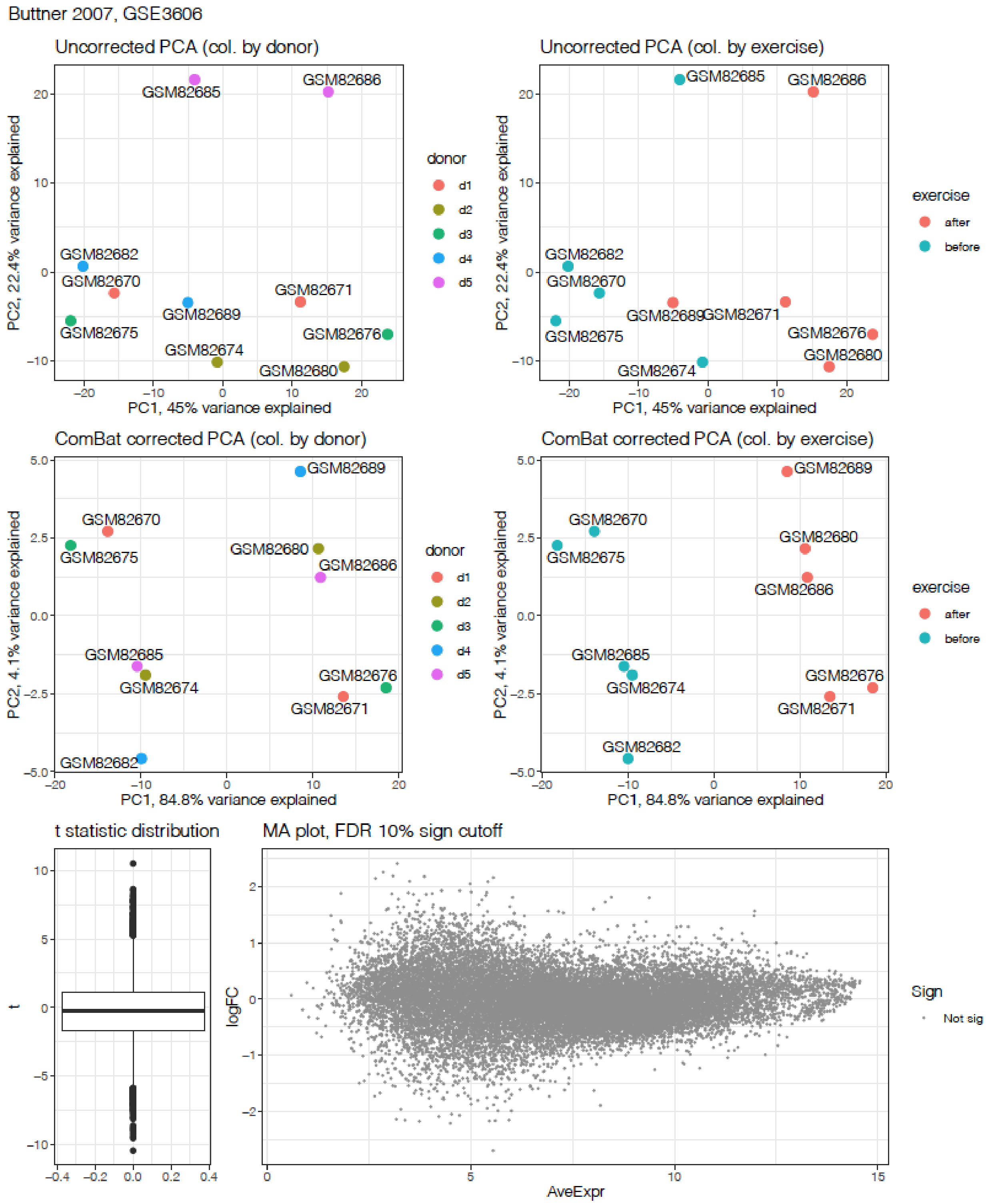


Figure S4. Microarray comparison of 5 adult athletes before and after exercise (Buttner 2007).

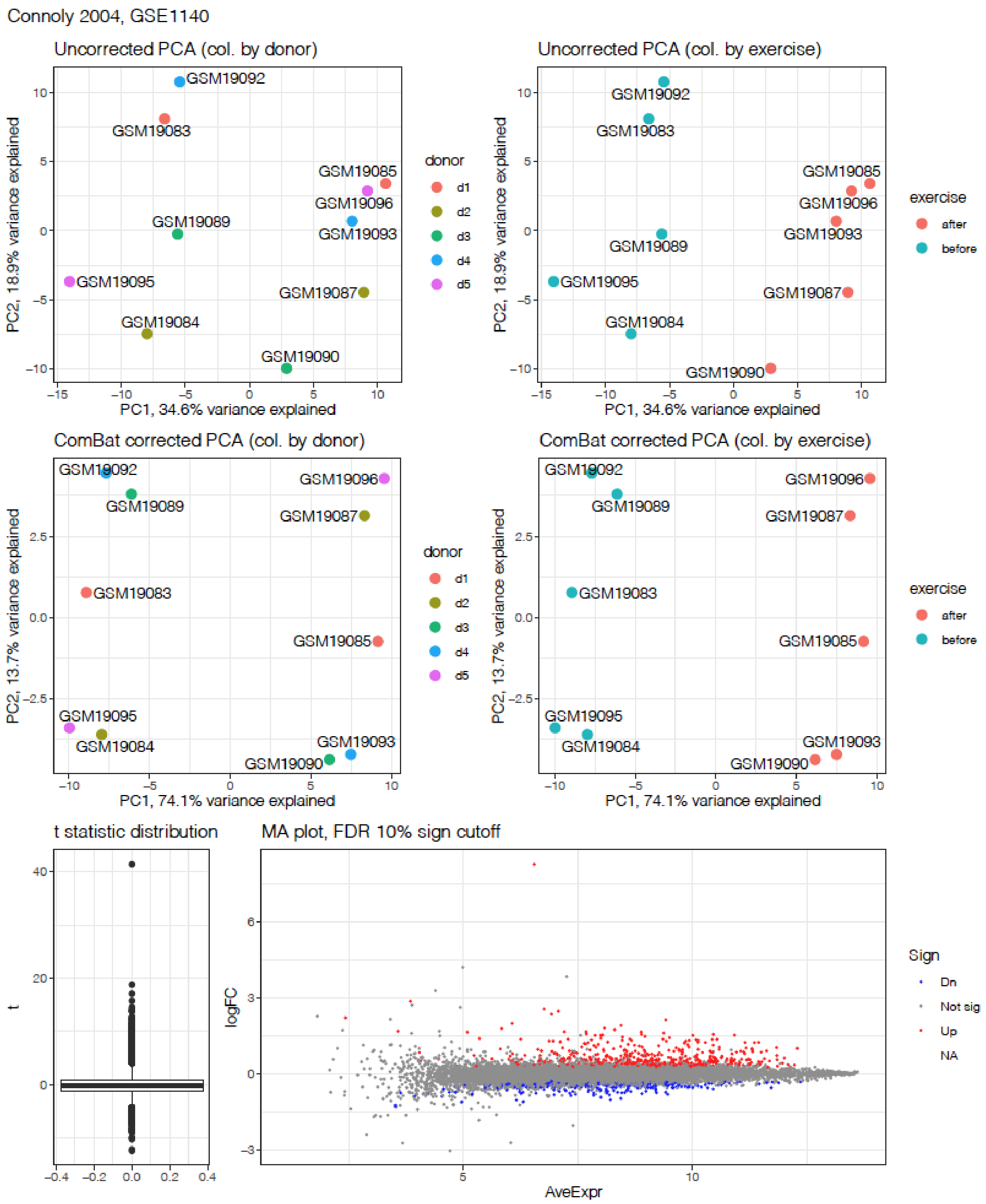


Figure S5. Microarray comparison of 5 adult athletes before and after exercise (Connoly 2004).

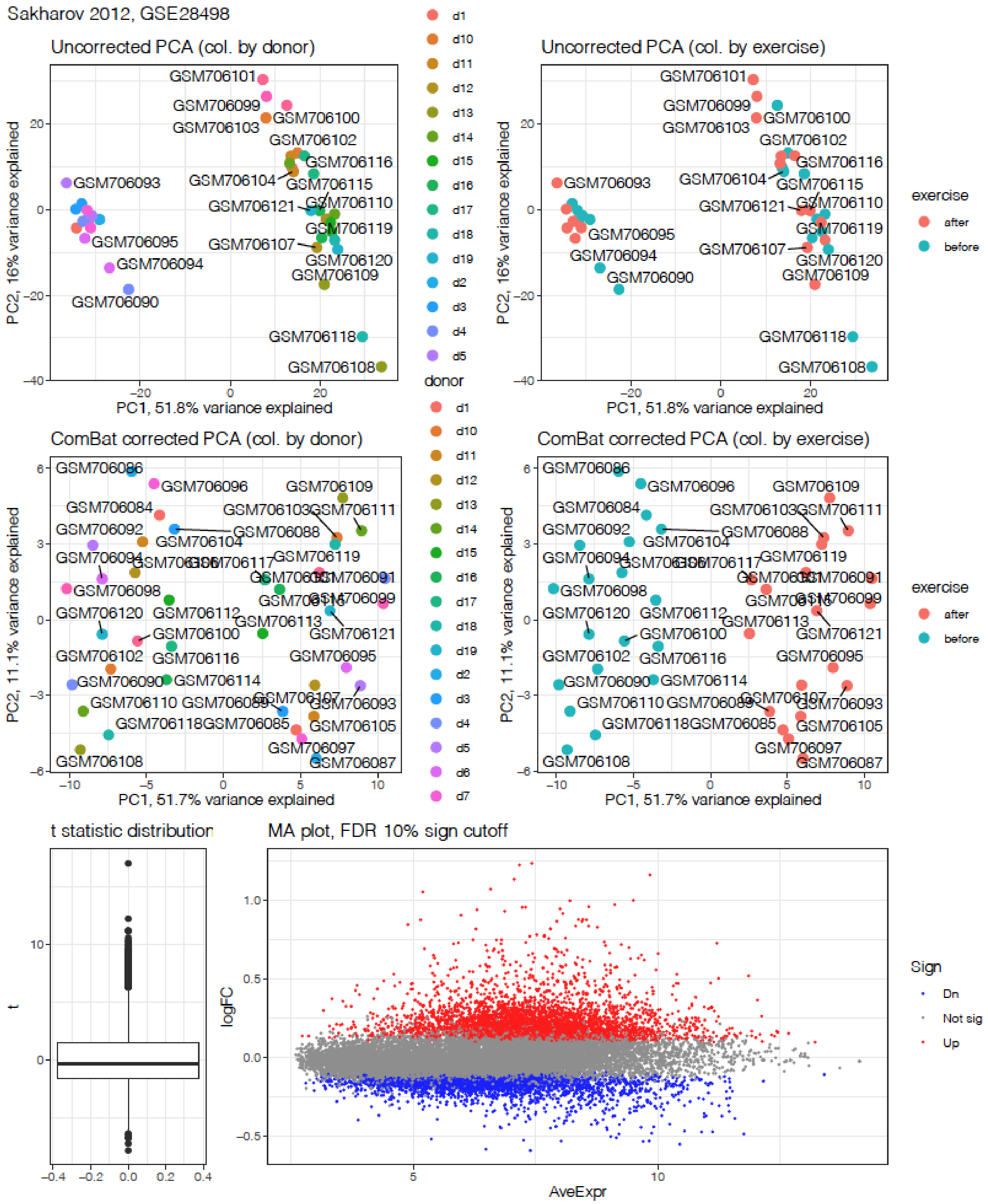


Figure S6. Microarray comparison of 19 adult athletes before and after exercise (Sakharov 2012).

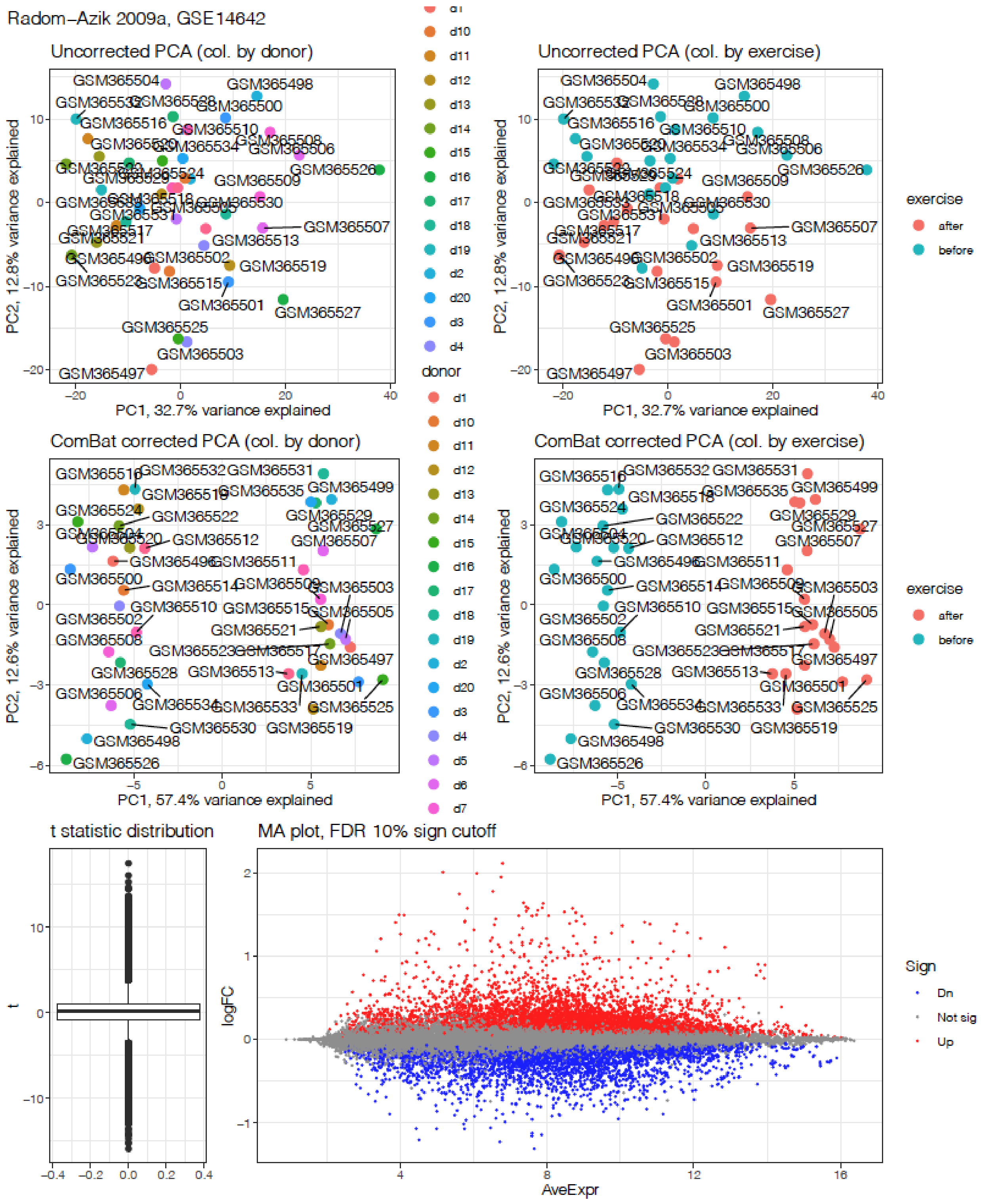


Figure S7. Microarray comparison of 20 teenage athletes before and after exercise (Radom-Azik 2009a).

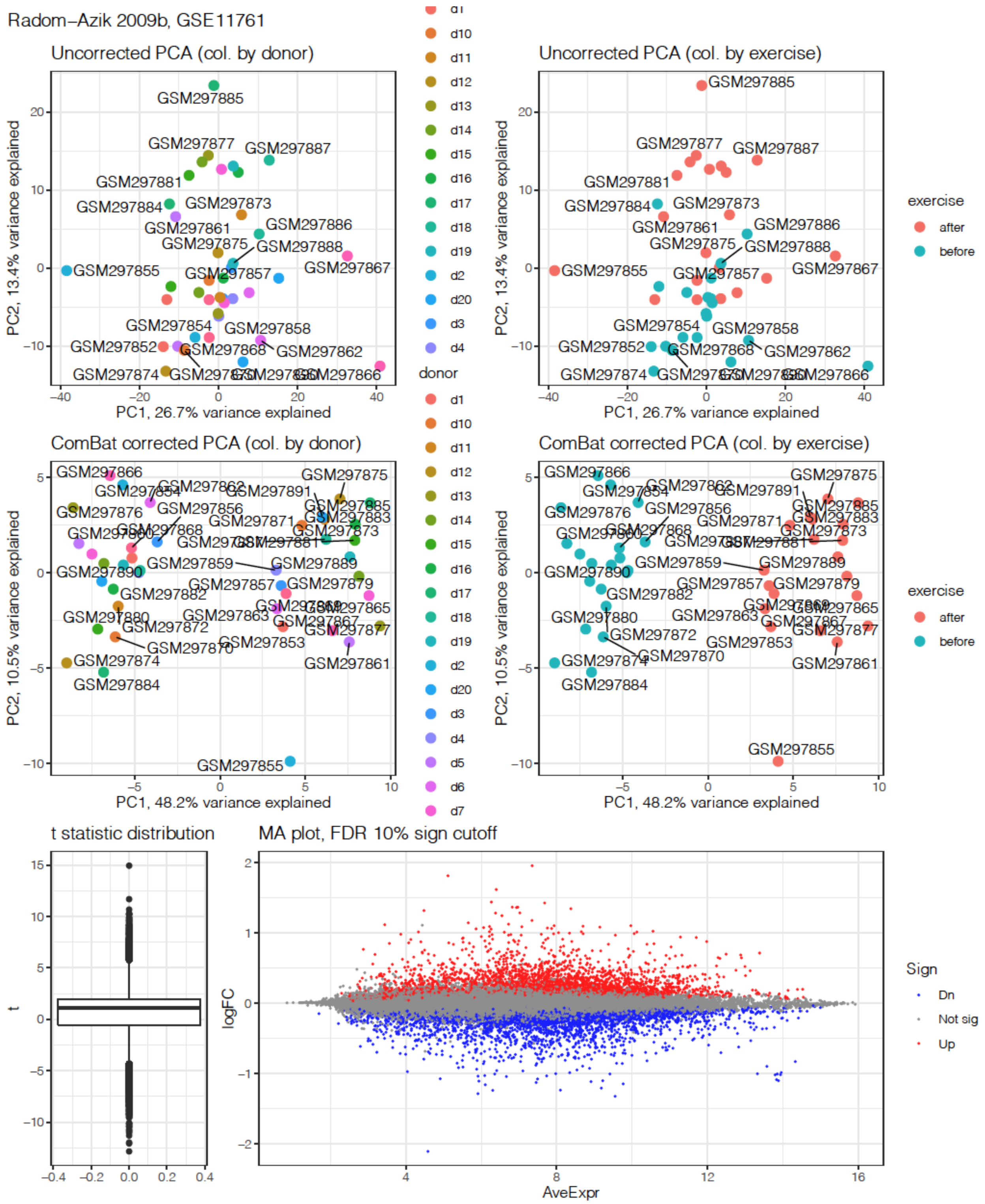


Figure S8. Microarray comparison of 20 teenage athletes before and after exercise (Radom-Azik 2009b).

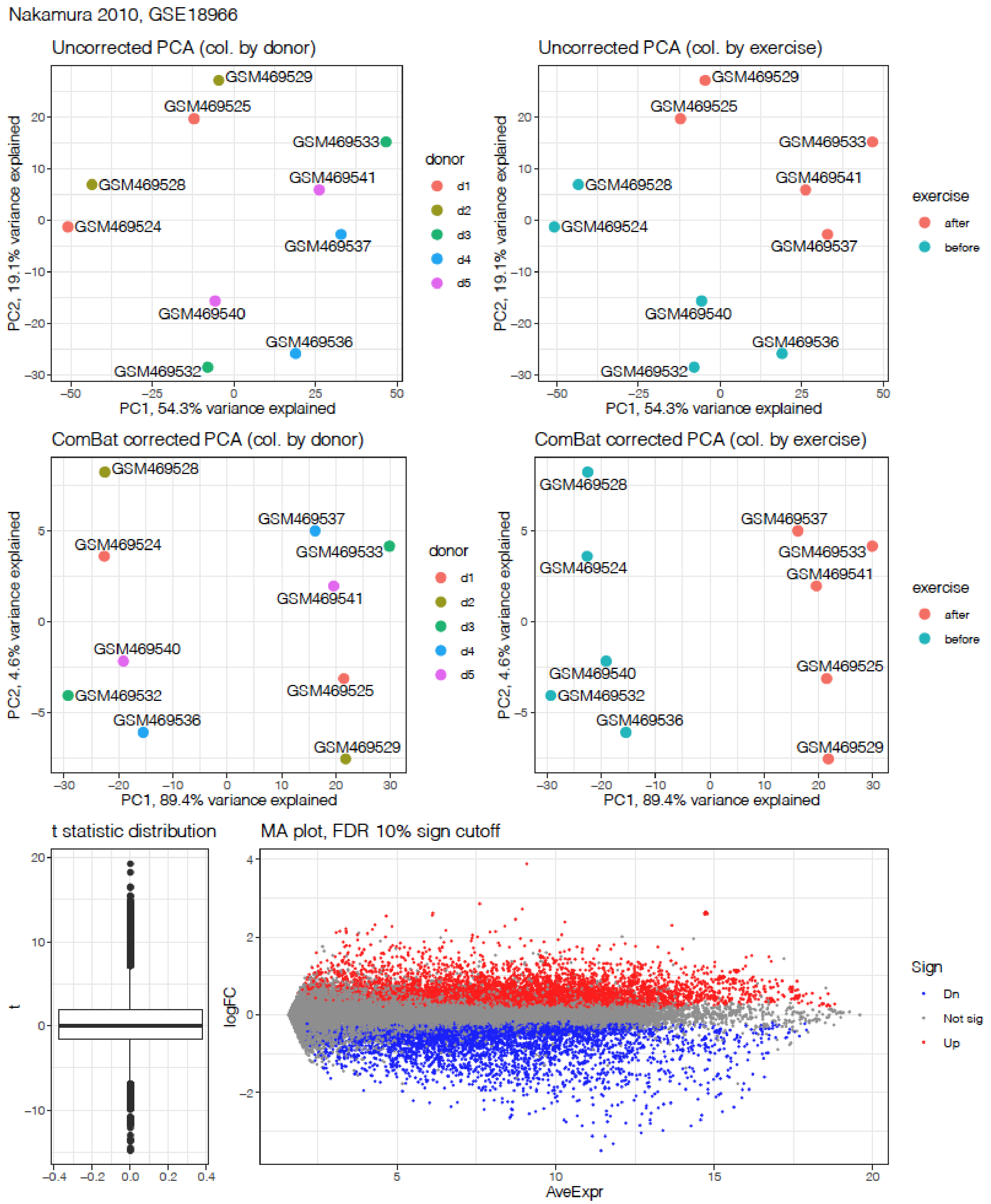


Figure S9. Microarray comparison of 5 adult athletes before and after exercise (Nakamura 2010).
